# Combining structured and unstructured data for predictive models: a deep learning approach

**DOI:** 10.1101/2020.08.10.20172122

**Authors:** Dongdong Zhang, Changchang Yin, Jucheng Zeng, Xiaohui Yuan, Ping Zhang

## Abstract

**Background:** The broad adoption of Electronic Health Records (EHRs) provides great opportunities to conduct health care research and solve various clinical problems in medicine. With recent advances and success, methods based on machine learning and deep learning have become increasingly popular in medical informatics. However, while many research studies utilize temporal structured data on predictive modeling, they typically neglect potentially valuable information in unstructured clinical notes. Integrating heterogeneous data types across EHRs through deep learning techniques may help improve the performance of prediction models.

**Methods:** In this research, we proposed 2 general-purpose multi-modal neural network architectures to enhance patient representation learning by combining sequential unstructured notes with structured data. The proposed fusion models leverage document embeddings for the representation of long clinical note documents and either convolutional neural network or long short-term memory networks to model the sequential clinical notes and temporal signals, and one-hot encoding for static information representation. The concatenated representation is the final patient representation which is used to make predictions.

**Results:** We evaluate the performance of proposed models on 3 risk prediction tasks (i.e., in-hospital mortality, 30-day hospital readmission, and long length of stay prediction) using derived data from the publicly available Medical Information Mart for Intensive Care III dataset. Our results show that by combining unstructured clinical notes with structured data, the proposed models outperform other models that utilize either unstructured notes or structured data only.

**Conclusions:** The proposed fusion models learn better patient representation by combining structured and unstructured data. Integrating heterogeneous data types across EHRs helps improve the performance of prediction models and reduce errors.

**Availability:** The code for this paper is available at: https://github.com/onlyzdd/clinical-fusion.

## Background

Electronic Health Records (EHRs) are longitudinal electronic records of patients’ health information, including structured data (patient demographics, vital signs, lab tests, etc.) and unstructured data (clinical notes and reports). In the United States, for example, over 30 million patients visit hospitals each year, and the percent of non-Federal acute care hospitals with the adoption of at least a Basic EHR system increased from 9.4% to 83.8% over the 7 years between 2008 and 2015 [1]. The broad adoption of EHRs provides unprecedented opportunities for data mining and machine learning researchers to conduct health care research.

With recent advances and success, machine learning and deep learning-based approaches have become increasingly popular in health care and shown great promise in extracting insights from EHRs. Accurately predicting clinical outcomes, such as mortality and readmission prediction, can help improve health care and reduce cost. Traditionally, some knowledge-driven scores are used to estimate the risk of clinical outcomes. For example, SAPS scores [2] and APACHE IV [3] are used to identify patients at high risk of mortality; LACE Index [4] and HOSPITAL Score [5] are used to evaluate hospital readmission risk. Recently, lots of research studies have been conducted for these prediction tasks based on EHRs using machine learning and deep learning techniques. Caruana [6] predicts hospital readmission using traditional logistic regression and random forest models. Tang [7] shows that recurrent neural networks using temporal physiologic features from EHRs provide additional benefits in mortality prediction. Rajkomar [8] combines 3 deep learning models and develops an ensemble model to predict hospital readmission and long length of stay. Besides, Min [9] compared different types of machine learning models for predicting the readmission risk of Chronic Obstructive Pulmonary Disease patients. Two benchmarks studies [10, 11] show that deep learning models consistently outperform all the other approaches over several clinical prediction tasks. In addition to structured EHR data such as vital signs and lab tests, unstructured data also offers promise in predictive modeling [12, 13]. Boag [14] explores several representations of clinical notes and their effectiveness on downstream tasks. Liu’s model [15] forecasts the onset of 3 kinds of diseases using medical notes. Sushil [16] utilizes a stacked denoised autoencoder and a paragraph vector model to learn generalized patient representation directly from clinical notes and the learned representation is used to predict mortality.

However, most of the previous works focused on prediction modeling by utilizing either structured data or unstructured clinical notes and few of them pay enough attention to combining structured data and unstructured clinical notes. Integrating heterogeneous data types across EHRs (unstructured clinical notes, time-series clinical signals, static information, etc.) presents new challenges in EHRs modeling but may offer new potentials.

Recently, some works [15, 17] extract structured data as text features such as medical named entities and numerical lab tests from clinical notes and then combine them with clinical notes to improve downstream tasks. However, their approaches are domain-specific and cannot easily be transferred to other domains. Besides, their structured data are extracted from clinical notes and may introduce errors compared to original signals.

In this paper, we aim at combining structured data and unstructured text directly through deep learning techniques for clinical risk predictions. Deep learning methods have made great progress in many areas [18] such as computer vision [19], speech recognition [20] and natural language processing [21] since 2012. The flexibility of deep neural networks makes it well-suited for the data fusion problem of combining unstructured clinical notes and structured data. Here, we propose 2 multi-modal neural network architectures learn patient representation and the patient representation is then used to predict patient outcomes. The proposed multi-modal neural network architectures are purpose-general and can be applied to other domains without effort.

To summarize, the contributions of our work are:

- We propose 2 general-purpose fusion models to combine temporal signals and clinical text which lead to better performance on 3 prediction tasks.
- We examine the capability of unstructured clinical text in predictive modeling.
- We present benchmark results of in-hospital mortality, 30-day readmission and long length of stay prediction tasks. We show that deep learning models consistently outperform baseline machine learning models.
- We compare and analyze the running time of proposed fusion models and baseline models.

## Methods

In this section, we describe the dataset, patient features, predictive tasks, and proposed general-purpose neural network architectures for combining unstructured data and structured data using deep learning techniques.

### Dataset description

Medical Information Mart for Intensive Care III (MIMIC-III) [22] is a publicly available critical care database maintained by the Massachusetts Institute of Technology (MIT)’s Laboratory for Computational Physiology. MIMIC-III comprises deidentified health-related data associated with over forty thousand patients who stayed in critical care units of the Beth Israel Deaconess Medical Center (BIDMC) between 2001 and 2012. This database includes patient health information such as demographics, vital signs, lab test results, medications, diagnosis codes, as well as clinical notes.

### Patient features

Patient features consist of features from both structured data (static information and temporal signals) and unstructured data (clinical text). In this part, we describe the patient features that are utilized by our model and some data preprocessing details.

#### Static information

Static information refers to demographic information and admission-related information in this study. For demographic information, patient’s age, gender, marital status, ethnicity, and insurance information are considered. Only adult patients are enrolled in this study. Hence, age was split into 5 groups (18, 25), (25, 45), (45, 65), (65, 89), (89,). For admission-related information, admission type is included as features.

#### Temporal signals

For temporal signals, we consider 7 frequently sampled vital signs: heart rate, systolic blood pressure (SysBP), diastolic blood pressure (DiasBP), mean arterial blood pressure (MeanBP), respiratory rate, temperature, SpO2; and 19 common lab tests: anion gap, albumin, bands, bicarbonate, bilirubin, creatinine, chloride, glucose, hematocrit, hemoglobin, lactate, platelet, potassium, partial thromboplastin time (PTT), international normalized ratio (INR), prothrombin time (PT), sodium, blood urea nitrogen (BUN), white blood cell count (WBC). Statistics of temporal signals are shown in Table 1. Additional statistics of temporal signals are provided in Supplementary Materials. After feature selection, we extract values of these time-series features up to the first 24 hours of each hospital admission. For each temporal signal, average is used to represent the signal at each timestep (hour). Then, each temporal variable was normalized using min-max normalization. To handle missing values, we simply use “0” to impute [23].

**Table 1.**
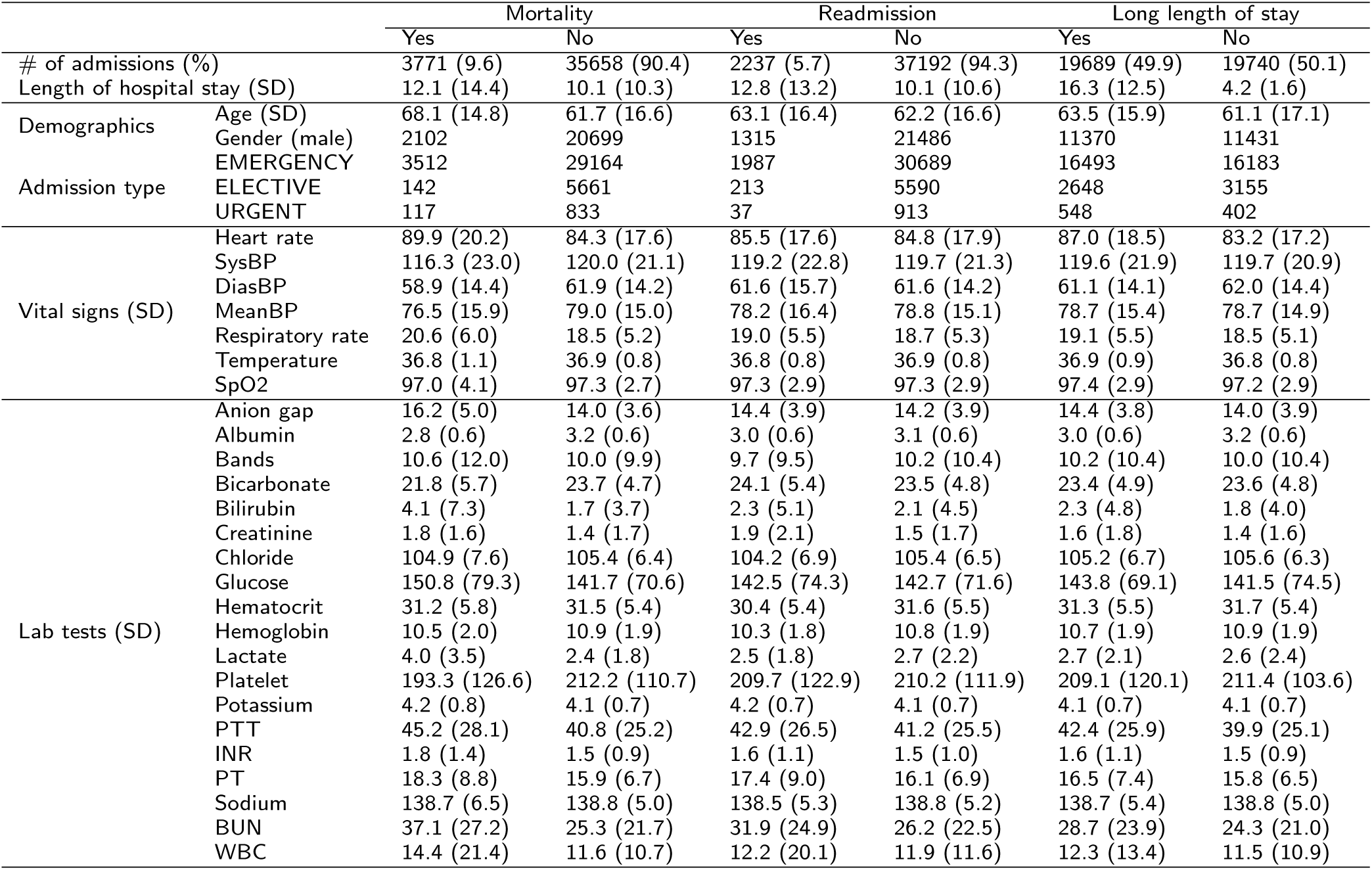
Label statistics and characteristics of 3 prediction tasks. SD represents standard deviation.

#### Sequential clinical notes

In addition to the aforementioned types of structured data, we also incorporate sequential unstructured notes, which contain a vast wealth of knowledge and insight that can be utilized for predictive models using Natural Language Processing (NLP). In this study, we considered *Nursing*, *Nursing/Other*, *Physician*, and *Radiology* notes, because these kinds of notes are in the majority of clinical notes and are frequently recorded in MIMIC-III database. We only extract the first 24 hours’ notes for each admission to enable early prediction of outcomes.

### Predictive tasks

Here, 3 benchmark prediction tasks are adopted which are crucial in clinical data problems and have been well studied in the medical community [7, 8, 24, 25, 26, 27, 28].

#### In-hospital mortality prediction

Mortality prediction is recognized as one of the primary outcomes of interest. The overall aim of this task is to predict whether a patient passes away during the hospital stay. This task is formulated as a binary classification problem, where the label indicates the occurrence of a death event. To evaluate the performance, we report the F1-score (F1), area under the receiver operating characteristic curve (AUROC), and area under the precision-recall curve (AUPRC). AUROC is the main metric.

#### Long length of stay prediction

Length of stay is defined as the time interval between hospital admission and discharge. In the second task, we predict a long length of stay whether a length of stay is more than 7 days [8, 28]. Long length of stay prediction is important for hospital management. This task is formulated as a binary classification problem with the same metrics of the mortality prediction task.

#### Hospital readmission prediction

Hospital readmission refers to unplanned hospital admissions within 30 days following the initial discharge. Hospital readmission has received much attention because of its No impacts on healthcare systems’ budgets. In the US, for example, roughly 2 million hospital readmissions each year costs Medicare 27 billion dollars, of which 17 billion dollars are potentially avoidable [29]. Reducing preventable hospital readmissions represents an opportunity to improve health care, lower costs, and increase patient satisfaction. Predicting unplanned hospital readmission is a binary classification problem with the same metrics as the in hospital mortality prediction task.

### Neural Network Architecture

In this part, we present 2 neural network architectures for combining clinical structured data with sequential clinical notes. The overview of the proposed models, namely Fusion-CNN and Fusion-LSTM, are illustrated in Figure 1 and Figure 2. Each model mainly consists of 5 parts, static information encoder, temporal signals embedding, sequential notes representation, patient representation, and output layer. Fusion-CNN is based on convolutional neural networks (CNN) and Fusion-LSTM is based on long-short term memory (LSTM) networks. The 2 models have common in model inputs and outputs but differ in the way how they model the temporal information.

**Figure 1.**
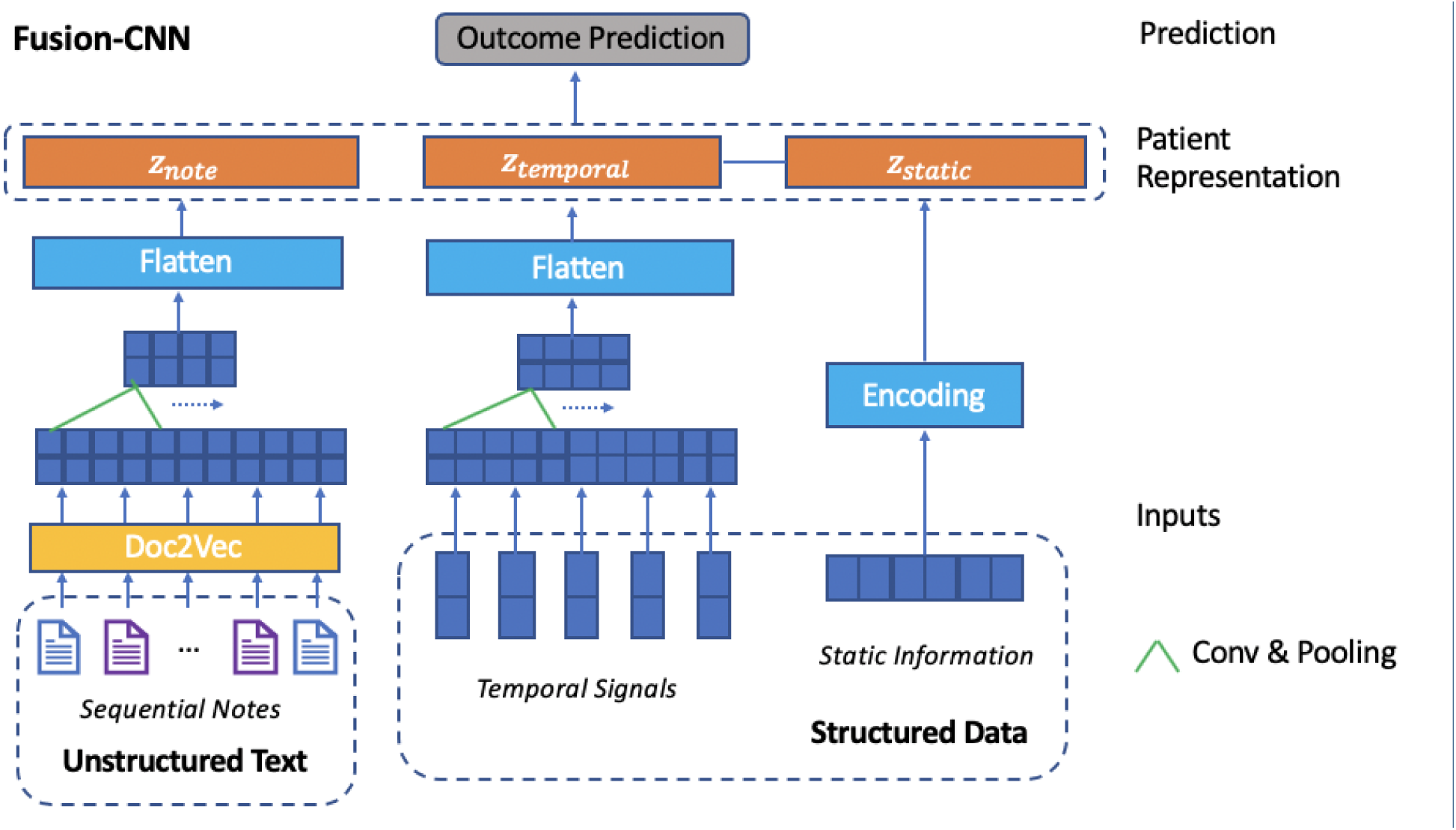
Architecture of CNN-based Fusion-CNN. Fusion-CNN uses document embeddings, 2-layer CNN and max-pooling to model sequential clinical notes. Similarly, 2-layer CNN and max-pooling are used to model temporal signals. The final patient representation is the concatenation of the latent representation of sequential clinical notes, temporal signals, and the static information vector. Then the final patient representation is passed to output layers to make predictions.

#### Static information encoder

The static categorical features including patient demographics and admission-related information are encoded as one-hot vectors through the static information encoder. The output of the encoder is *z_static_* = *[z_demo_; z_adm_*] with size *d_static_ = d_demo_* + *d_adm_* and *z_static_* is part of patient representation.

#### Temporal signals representation

In this part, Fusion-CNN and Fusion-LSTM leverage different techniques to model temporal signals. The learned vector for temporal signals representation is *z_temporal_* with size of *d_temporal_*.

**Fusion-CNN** Convolutional neural networks (CNNs) can automatically learn the features through convolution and pooling operations and can be used for time-series modeling. Fusion-CNN uses 2-layer convolution and max-pooling to extract deep features of temporal signals as shown in Figure 1.

**Fusion-LSTM** Recurrent neural networks (RNNs) are considered since RNN models have achieved great success in sequences and time series data modeling. However, RNNs with simple activations suffer from vanishing gradients. Long short-term memory (LSTM) neural networks are a type of RNNs that can learn and remember long sequences of input data. 2-layer LSTM is utilized in Fusion-LSTM model to learn the representations of temporal signals as shown in Figure 2. To prevent the model from overfitting, dropout on nonrecurrent connections is applied between RNN layers and before outputs.

**Figure 2.**
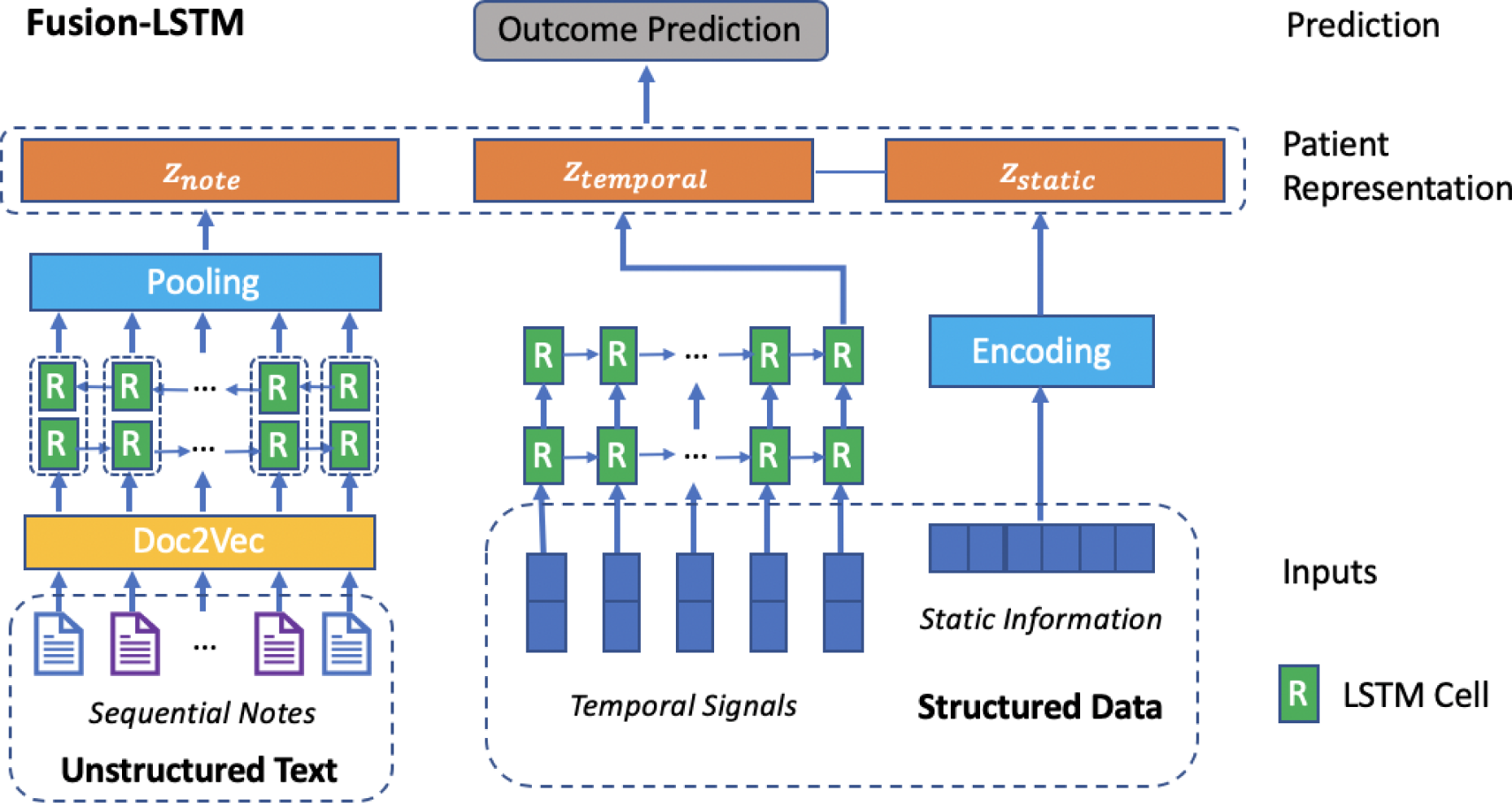
Architecture of LSTM-based Fusion-LSTM. Fusion-LSTM uses document embeddings, a BiLSTM layer, and a max-pooling layer to model sequential clinical notes. 2-layer LSTMs are used to model temporal signals. The concatenated patient representation is passed to output layers to make predictions.

#### Sequential notes representation

Word embedding is a popular technique in natural language processing that is used to map words or phrases from vocabulary to a corresponding vector of continuous values. However, directly modeling sequential notes using word embeddings and deep learning can be time-consuming and may not be practical since clinical notes are usually very long and involve multiple timestamps. To solve this problem, we present the sequential notes representation component based on document embeddings. Here, we utilize paragraph vector (aka. Doc2Vec) [30] to learn the embedding of each clinical note. Time-series document embeddings are inputs to Fusion-CNN and Fusion-LSTM as shown in Figure 1 and Figure 2. The sequential notes representation component produces *z_note_* with size of *d_note_* as the latent representation of sequential notes.

**Fusion-CNN** As shown in Figure 1, the sequential notes representation part of Fusion-CNN model is made up of document embeddings, a series of convolutional layers and max-pooling layers, and a flatten layer. Document embedding inputs are passed to these convolutional layers and max-pooling layers. The flatten layer takes the output of the max-pooling layer as input and outputs the final text representation.

**Fusion-LSTM** model is demonstrated in Figure 2, the sequential notes representation part is made up of document embeddings, a BiLSTM (bidirectional LSTM) layer, and a max-pooling layer. The document embedding inputs are passed to the BiL-STM layer. The BiLSTM layer concatenates the outputs (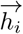, 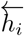) from 2 hidden layers of opposite direction to the same output 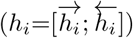 and can capture long term dependencies in sequential text data. The max-pooling layer takes the hidden states of the BiLSTM layer as input and outputs the final text representation.

#### Patient representation

The final patient representation *z* is obtained by concatenating the representations of clinical text, temporal signals, along with static information. The representation of each patient is *z_p_ =* [*z_static_*; *z_temporal_; z_text_*], the size of this vector is *d_static_* + *d_temporal_* + *d_text_*. The patient representation is then fed to a final output layer to make predictions.

#### Output layer

The output layer takes patient representation as input and makes predictions. For each patient representation *z_p_*, we have a task-specific target *y. y* ∊ {0, 1} is a single binary label indicating whether the in-hospital mortality, 30-day readmission, or long length of stay event occurs.

For each prediction task, the output layer receives an instance of patient representation *z_p_* as input and tries to predict the ground truth *y*. For binary classification tasks, the output layer is:

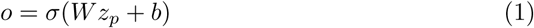

The *W* matrices and *b* vectors are the trainable parameters, *σ* represents a sigmoid activation function.

For each of these 3 tasks, the loss functions is defined as the binary cross entropy loss:

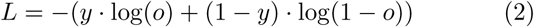

## Results and Discussion

### Experiment setup

#### Cohort preparation

Based on the MIMIC-III dataset, we evaluated our proposed models on 3 predictive tasks (i.e. in-hospital mortality prediction, 30-day readmission prediction, and long length of stay prediction). To build corresponding cohorts, we first removed all patients whose age <18 years old and all hospital admissions whose length of stay is less than 1 day. Besides, patients without any records of required temporal signals and clinical notes were removed. In total, 39,429 unique admissions are eligible for prediction tasks. Label statistics and characteristics of 3 prediction tasks are provided in Table 1.

#### Implementation details

In this part, we describe the implementation details. We train the unsupervised Doc2Vec model on the training set to obtain the document-level embeddings for each note using the popular Gensim toolkit [31]. We use PV-DBOW as the training algorithm, number of training epochs as 30, initial learning rate as 0.025, learning rate decay as 0.0002, and dimension of vectors as 200 to train.

We implement baseline models (i.e., logistic regression and random forest) with scikit-learn [32]. Deep learning models are implemented using PyTorch [33]. All deep learning models are trained with Adam optimizer with a learning rate of 0.0001 and ReLU as the activation function. The batch size is chosen as 64 and the max epoch number is set to 50.

For evaluation, 70% of the data are used for training, and 10% for validation, 20% for testing. For binary classification tasks, AUROC is used as the main metric. Besides, we report F1 score, and AUPRC to aid the interpretation of AUROC for imbalanced datasets.

#### Baselines

We compared our model with the following baseline methods: logistic regression (LR), random forest (RF). Because these standard machine learning methods cannot work directly with temporal sequences, the element-wise mean vector across sequential notes and aggregations (i.e. mean value, minimum value, maximum value, standard deviation, and count of observations) of temporal signals are used as model inputs.

#### Ablation study

To evaluate the contribution of different components and gain a better understanding of the proposed fusion model’s behavior, an ablation study is adopted and we have conducted extensive experiments on different models. Let U, T, S denote the unstructured clinical notes, temporal signals, and static information.

## Results

In this section, we report the performance of proposed models on 3 prediction tasks. The results are shown in Table 2, Table ??, and Table 6. Each reported performance metric is the average score of 5 runs with different data splits. To measure the uncertainty of a trained model’s performance, we calculated 95% confidence interval using t-distribution and the results are reported. Besides, to better compare model performances on each task, we performed statistical testing and calculated P-value of AUROC score across various models using statistical t-testingP-value matrix of AUROC scores on in-hospital mortality prediction, long length of stay prediction, 30-day readmission prediction tasks are shown in Table 3, Table 5, and Table 7. In summary, the results shows significant improvements and it matches our expectations: (1) Deep learning models outperformed traditional machine learning models by comparing the performances of different models on the same model inputs. (2) Models could make more accurate predictions by combining unstructured text and structured data.

**Table 2.**
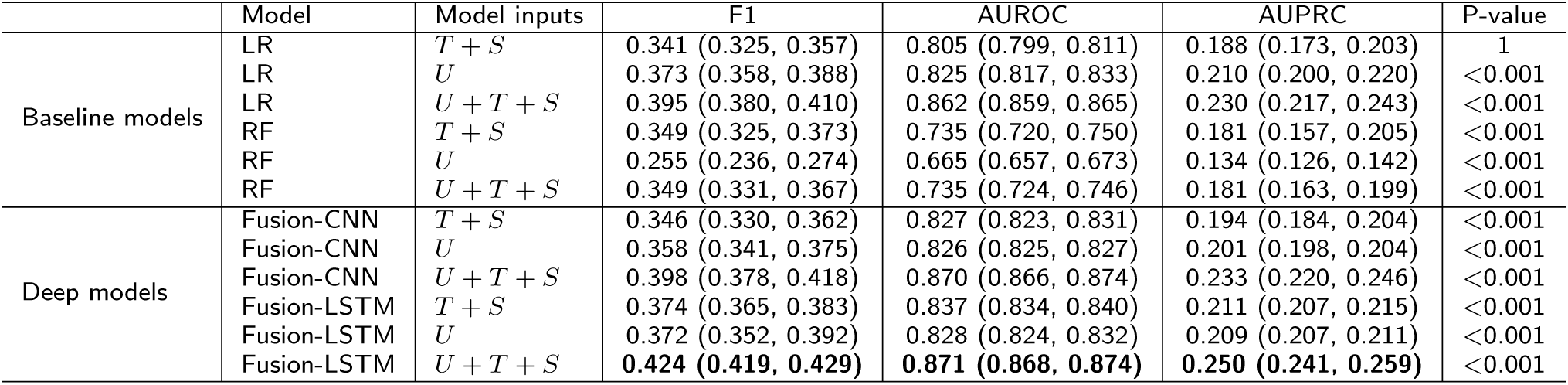
In-hospital mortality prediction on MIMIC-III. *U*, *T*, *S* represents unstructured data, temporal signals, and static information respectively.

**Table 3.**
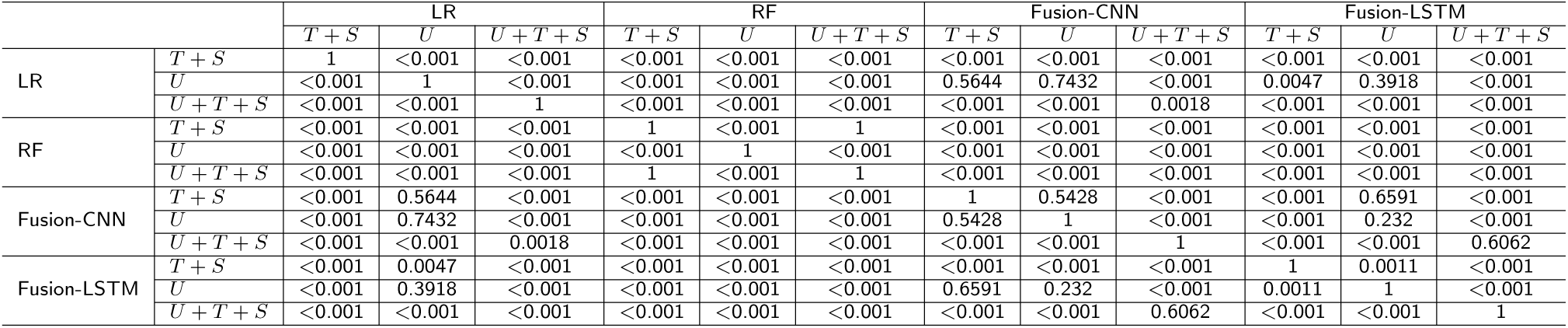
P-value matrix of various model performances (AUROC) for in-hospital mortality prediction. *U*, *T*, *S* represents unstructured data, temporal signals, and static information respectively.

### In-hospital mortality prediction

Table 2 shows the performance of various models on the in-hospital mortality prediction task. From Table 2 and Table 3, deep models outperformed baseline models. We speculate the main reasons why deep models work better are two-fold: (1) Deep models can automatically learn better patient representations as the network grows deeper and yield more accurate predictions. (2) Deep models can capture temporal information and local patterns, while logistic regression and random forest simply aggregate time-series features and hence suffer from information loss.

For each kind of classifier, the performance of classifier trained on all data (*U*+*T*+*S*) is significantly higher than that trained on either structured data (*T* + *S*) or unstructured data (*U*) only. Especially by considering unstructured text, the AUROC score of Fusion-CNN and Fusion-LSTM increased by 0.043 and 0.034 respectively. Structured data contains a patient’s vital signs and lab test results, while sequential notes provide the patient’s clinical history including diagnoses, medications, and so on. This observation implicitly explains why unstructured text and structured data can complement each other to some extent in predictive modeling which leads to performance improvement.

### Long length of stay prediction

Table 4 shows the performance measured by F1, AUROC, and AUPRC of different models on the long length of stay prediction. We observe that (1) Logistic regression serves as a very strong baseline while Fusion-CNN achieves a slightly higher F1 score and AUROC score than Fusion-LSTM. (2) By integrating multi-modal information, all models yield more accurate predictions and the improvement is significant as shown in Table 5.

**Table 4.** Long length of stay prediction on MIMIC-III. *U*, *T*, *S* represents unstructured data, temporal signals, and static information respectively.

|  | Model | Model inputs | F1 | AUROC | AUPRC | P-value |
| --- | --- | --- | --- | --- | --- | --- |
| Baseline models | LR | $T + S$ | 0.668 (0.658, 0.678) | 0.735 (0.732, 0.738) | 0.615 (0.611, 0.619) | 1 |
| | LR | $U$ | 0.686 (0.683, 0.689) | 0.736 (0.732, 0.740) | 0.614 (0.610, 0.618) | 0.5643 |
| | LR | $U + T + S$ | 0.703 (0.699, 0.707) | 0.773 (0.770, 0.776) | 0.642 (0.637, 0.647) | <0.001 |
| | RF | $T + S$ | 0.523 (0.462, 0.584) | 0.695 (0.689, 0.701) | 0.586 (0.577, 0.595) | <0.001 |
| | RF | $U$ | 0.568 (0.479, 0.657) | 0.651 (0.642, 0.660) | 0.559 (0.553, 0.565) | <0.001 |
| | RF | $U + T + S$ | 0.537 (0.533, 0.541) | 0.718 (0.714, 0.722) | 0.597 (0.591, 0.603) | <0.001 |
| Deep models | Fusion-CNN | $T + S$ | 0.674 (0.667, 0.681) | 0.748 (0.745, 0.751) | 0.640 (0.635, 0.645) | <0.001 |
| | Fusion-CNN | $U$ | 0.695 (0.683, 0.707) | 0.742 (0.741, 0.743) | 0.635 (0.632, 0.638) | <0.001 |
| | Fusion-CNN | $U + T + S$ | **0.725 (0.718, 0.732)** | **0.784 (0.781, 0.787)** | **0.662 (0.658, 0.666)** | <0.001 |
| | Fusion-LSTM | $T + S$ | 0.690 (0.684, 0.696) | 0.757 (0.756, 0.758) | 0.644 (0.643, 0.645) | <0.001 |
| | Fusion-LSTM | $U$ | 0.702 (0.697, 0.707) | 0.746 (0.745, 0.747) | 0.637 (0.634, 0.640) | <0.001 |
| | Fusion-LSTM | $U + T + S$ | 0.716 (0.711, 0.721) | 0.778 (0.776, 0.780) | 0.660 (0.657, 0.663) | <0.001 |

**Table 5.**
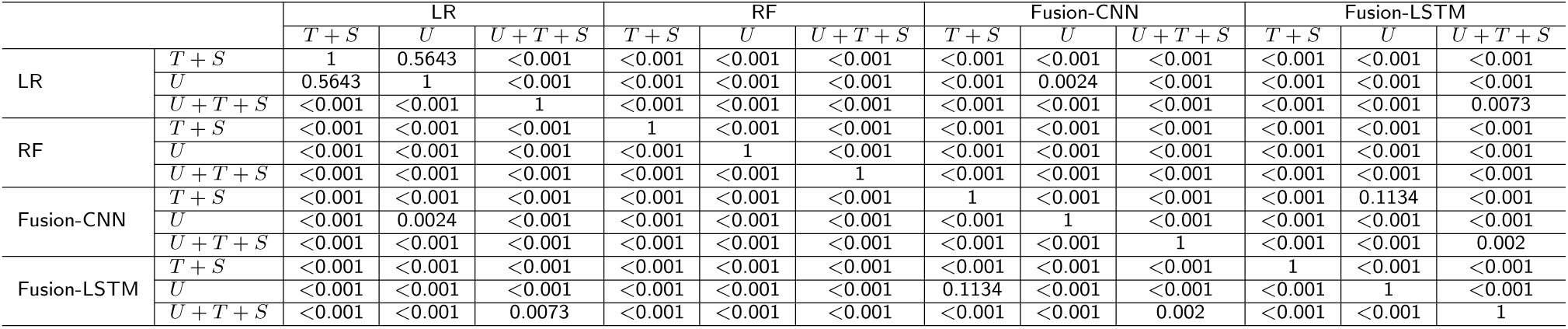
P-value matrix of various model performances (AUROC) for long length of stay prediction. *U*, *T*, *S* represents unstructured data, temporal signals, and static information respectively.

### Hospital readmission prediction

Table 6 and Table 7 summarize the results of various approaches of the hospital readmission task. For this task, logistic regression performed well but random forest performed badly. Fusion-CNN and Fusion-LSTM yielded comparably better predictions of AU-ROC score around 0.67. Incorporating clinical notes led to performance improvement for logistic regression, Fusion-CNN, and Fusion-LSTM. However, combining unstructured notes with structured data hurt the performance of random forest.

We noted the AUROC score for hospital readmission prediction is significantly lower than in-hospital mortality one which means readmission risk modeling is more complex and difficult compared to in hospital mortality prediction. This is probably because the given features are inadequate for building a good hospital readmission risk prediction model. Besides, we only used the first day’s data which is far away from patient discharge that may not be very helpful in readmission prediction modeling.

**Table 6.** 30-day readmission prediction on MIMIC-111. *U*, *T*, *S* represents unstructured data, temporal signals, and static information respectively.

|  | Model | Model inputs | F1 | AUROC | AUPRC | P-value |
| --- | --- | --- | --- | --- | --- | --- |
| Baseline models | LR | *T* + *S* | 0.144 (0.136, 0.152) | 0.649 (0.646, 0.652) | 0.071 (0.062, 0.080) | 1 |
|  | LR | *U* | 0.142 (0.133, 0.151) | 0.638 (0.634, 0.642) | 0.070 (0.056, 0.084) | <0.001 |
|  | LR | *U* + *T* + *S* | 0.144 (0.137, 0.151) | 0.660 (0.657, 0.663) | 0.072 (0.059, 0.085) | <0.001 |
|  | RF | *T* + *S* | 0.123 (0.113, 0.133) | 0.575 (0.559, 0.591) | 0.060 (0.054, 0.066) | <0.001 |
|  | RF | *U* | 0.117 (0.105, 0.129) | 0.557 (0.539, 0.575) | 0.059 (0.056, 0.062) | <0.001 |
|  | RF | *U* + *T* + *S* | 0.118 (0.111, 0.125) | 0.560 (0.543, 0.577) | 0.059 (0.056, 0.062) | <0.001 |
| Deep models | Fusion-CNN | *T* + *S* | 0.155 (0.146, 0.164) | 0.657 (0.650, 0.664) | 0.077 (0.073, 0.081) | 0.0208 |
|  | Fusion-CNN | *U* | 0.163 (0.160, 0.166) | 0.663 (0.660, 0.666) | 0.078 (0.077, 0.079) | <0.001 |
|  | Fusion-CNN | *U* + *T* + *S* | **0.164 (0.161, 0.167)** | 0.671 (0.668, 0.674) | **0.080 (0.076, 0.084)** | <0.001 |
|  | Fusion-LSTM | *T* + *S* | 0.149 (0.146, 0.152) | 0.653 (0.651, 0.655) | 0.074 (0.071, 0.077) | 0.0158 |
|  | Fusion-LSTM | *U* | 0.158 (0.154, 0.162) | 0.641 (0.635, 0.647) | 0.075 (0.072, 0.078) | 0.0076 |
|  | Fusion-LSTM | *U* + *T* + *S* | 0.160 (0.151, 0.169) | **0.674 (0.672, 0.676)** | 0.079 (0.076, 0.082) | <0.001 |

**Table 7.**
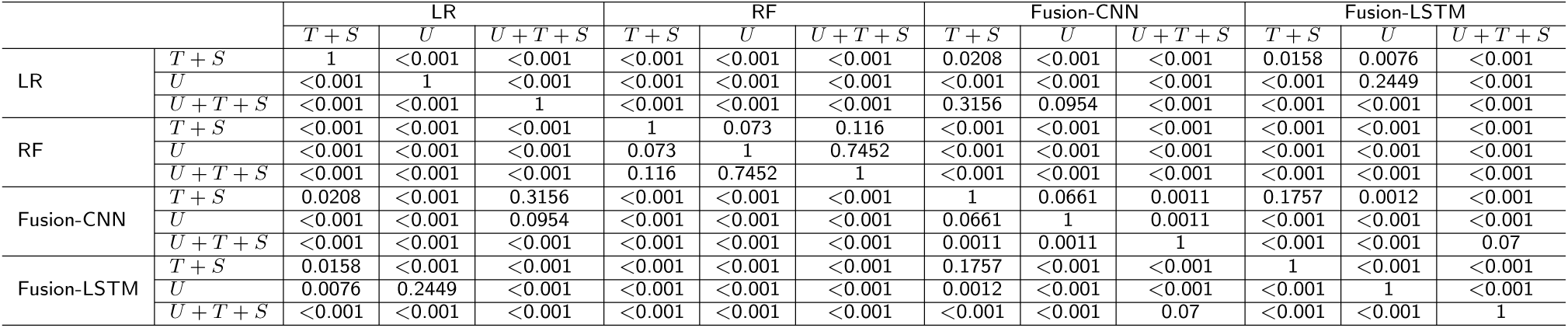
P-value matrix of various model performances (AUROC) for 30-day readmission prediction. *U*, *T*, *S* represents unstructured data, temporal signals, and static information respectively.

## Discussion

In this study, we examined proposed fusion models on 3 outcome prediction tasks, namely mortality prediction, long length of stay prediction, and readmission prediction. The results showed that deep fusion models (Fusion-CNN, Fusion-LSTM) outperformed baselines and yielded more accurate predictions by incorporating unstructured text.

In 3 tasks, logistic regression was a quite strong baseline and was consistently more useful than random forest. Deep models achieved the best performance for each task while training time of deep models is also acceptable as demonstrated in Figure 3. All experiments were performed on a 32-core Intel(R) Core(TM) i9-9960X CPU @ 3.10GHz machine with NVIDIA TITAN RTX GPU processor. For a fair comparison, we report the training time per epoch for Fusion-CNN and Fusion-LSTM.

**Figure 3.**
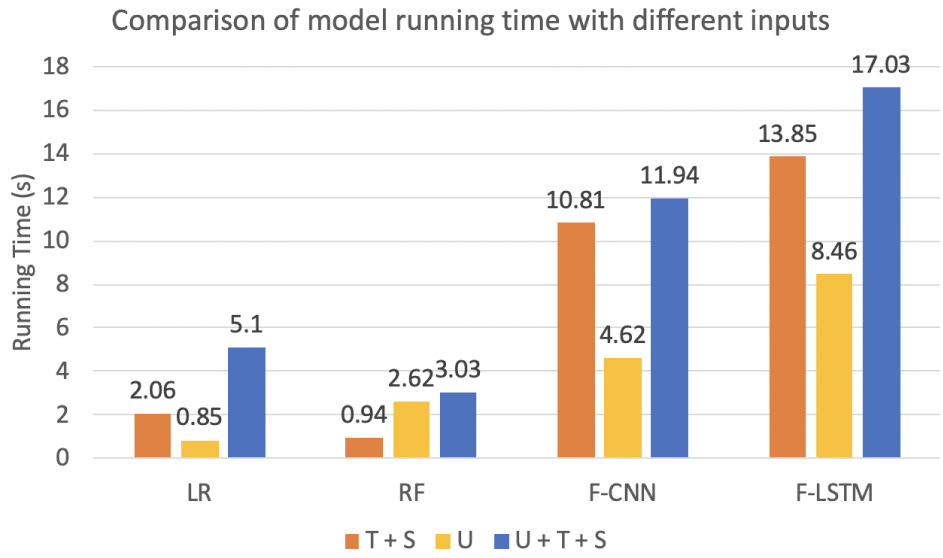
Comparison of model running time with different inputs.

## Conclusion

In this paper, we proposed 2 multi-modal deep neural networks that learn patient representation by combining unstructured clinical text and structured data. The 2 models make use of either LSTMs or CNNs to model temporal information. The proposed models are quite general data fusion methods and can be which mainly due to the learned patient representation consists of medication, diagnosis information from sequential unstructured notes, and vital signs, lab test results from structured data. In future work, we plan to apply the proposed fusion methods to more real-world applications.

## Data Availability

MIMIC-III database analyzed in the study is available on PhysioNet repository.

https://mimic.physionet.org/about/mimic

## Additional Files

Additional File 1 — Additional statistics.

Figure S1. Length of stay distribution of the processed MIMIC-III cohort.

Table S1. Statistics of collected vital signs and lab tests.

## Acknowledgements

Not applicable.

## Author’s contributions

PZ conceived the project. DZ and PZ developed the method. DZ conducted the experiments. DZ, CY, JZ, and PZ analyzed experimental results. DZ, CY, JZ, XY, and PZ wrote the manuscript. All authors read and approved the final manuscript.

## Funding

This project was funded in part under a grant with Lyntek Medical Technologies, Inc.

## Availability of data and materials

MIMIC-III database analyzed in the study is available on PhysioNet repository. The source code is provided for reproducing and is available at https://github.com/onlyzdd/clinical-fusion.

## Ethics approval and consent to participate

Not applicable.

## Consent for publication

Not applicable.

## Competing interests

PZ is the member of the editorial board of BMC Medical Informatics and Decision Making. The authors declare that they have no other competing interests.

## Notes

### Author Declarations

This study uses the MIMIC-III dataset. We are using the MIMIC IRB. This study was approved by the Institutional Review Boards of Beth Israel Deaconess Medical Center (Boston, MA, USA), the Massachusetts Institute of Technology (Cambridge, MA, USA). Requirement for individual patient consent was waived because the study did not impact clinical care and all protected health information was de-identified. De-identification was performed in compliance with Health Insurance Portability and Accountability Act (HIPAA) standards in order to facilitate public access to MIMIC-III. Deletion of protected health information (PHI) from structured data sources (e.g., database fields that provide patient name or date of birth) was straightforward.

## References

1. Henry J, Pylypchuk Y, Searcy T, Patel V. Adoption of electronic health record systems among US non-federal acute care hospitals: 2008-2015. ONC Data Brief. 2016;35:1–9.

2. Bisbal M, Jouve E, Papazian L, de Bourmont S, Perrin G, Eon B, et al. E_ectiveness of SAPS III to predict hospital mortality for post-cardiac arrest patients. Resuscitation. 2014;85(7):939–944.

3. Zimmerman JE, Kramer AA, McNair DS, Malila FM. Acute Physiology and Chronic Health Evaluation (APACHE) IV: hospital mortality assessment for today's critically ill patients. Critical care medicine. 2006;34(5):1297–1310.

4. van Walraven C, Dhalla IA, Bell C, Etchells E, Stiell IG, Zarnke K, et al. Derivation and validation of an index to predict early death or unplanned readmission after discharge from hospital to the community. Cmaj. 2010;182(6):551–557.

5. Donzé J, Aujesky D, Williams D, Schnipper JL. Potentially avoidable 30-day hospital readmissions in medical patients: derivation and validation of a prediction model. JAMA internal medicine. 2013;173(8):632–638.

6. Caruana R, Lou Y, Gehrke J, Koch P, Sturm M, Elhadad N. Intelligible models for healthcare: Predicting pneumonia risk and hospital 30-day readmission. In: Proceedings of the 21th ACM SIGKDD International Conference on Knowledge Discovery and Data Mining. ACM; 2015. p. 1721–1730.

7. Tang F, Xiao C, Wang F, Zhou J. Predictive modeling in urgent care: a comparative study of machine learning approaches. JAMIA Open. 2018;1(1):87–98.

8. Rajkomar A, Oren E, Chen K, Dai AM, Hajaj N, Hardt M, et al. Scalable and accurate deep learning with electronic health records. NPJ Digital Medicine. 2018;1(1):18.

9. Min X, Yu B, Wang F. Predictive modeling of the hospital readmission risk from patients' claims data using machine learning: a case study on COPD. Scientific reports. 2019;9(1):1–10.

10. Purushotham S, Meng C, Che Z, Liu Y. Benchmarking deep learning models on large healthcare datasets. Journal of biomedical informatics. 2018;83:112–134.

11. Harutyunyan H, Khachatrian H, Kale DC, Ver Steeg G, Galstyan A. Multitask learning and benchmarking with clinical time series data. Scienti_c data. 2019;6(1):96.

12. Grnarova P, Schmidt F, Hyland SL, Eickhoff C. Neural document embeddings for intensive care patient mortality prediction. arXiv preprint arXiv:161200467. 2016;.

13. Ghassemi M, Naumann T, Joshi R, Rumshisky A. Topic models for mortality modeling in intensive care units. In: ICML machine learning for clinical data analysis workshop; 2012. p. 1–4.

14. Boag W, Doss D, Naumann T, Szolovits P. What's in a note? Unpacking predictive value in clinical note representations. AMIA Summits on Translational Science Proceedings. 2018;2018:26.

15. Liu J, Zhang Z, Razavian N. Deep EHR: Chronic Disease Prediction Using Medical Notes. Journal of Machine Learning Research (JMLR). 2018;.

16. Sushil M, Šuster S, Luyckx K, Daelemans W. Patient representation learning and interpretable evaluation using clinical notes. Journal of biomedical informatics. 2018;84:103–113.

17. Jin M, Bahadori MT, Colak A, Bhatia P, Celikkaya B, Bhakta R, et al. Improving hospital mortality prediction with medical named entities and multimodal learning. arXiv preprint arXiv:181112276. 2018;.

18. LeCun Y, Bengio Y, Hinton G. Deep learning. nature. 2015;521(7553):436–444.

19. Wan J, Wang D, Hoi SCH, Wu P, Zhu J, Zhang Y, et al. Deep learning for content-based image retrieval: A comprehensive study. In: Proceedings of the 22nd ACM international conference on Multimedia. ACM; 2014. p. 157–166.

20. Deng L, Hinton G, Kingsbury B. New types of deep neural network learning for speech recognition and related applications: An overview. In: 2013 IEEE International Conference on Acoustics, Speech and Signal Processing. IEEE; 2013. p. 8599–8603.

21. Collobert R, Weston J. A united architecture for natural language processing: Deep neural networks with multitask learning. In: Proceedings of the 25th international conference on Machine learning. ACM; 2008. p. 160–167.

22. Johnson AE, Pollard TJ, Shen L, Li-wei HL, Feng M, Ghassemi M, et al. MIMIC-III, a freely accessible critical care database. Scientific data. 2016;3:160035.

23. Hu Z, Melton GB, Arsoniadis EG, Wang Y, Kwaan MR, Simon GJ. Strategies for handling missing clinical data for automated surgical site infection detection from the electronic health record. Journal of biomedical informatics. 2017;68:112–120.

24. Luo YF, Rumshisky A. Interpretable topic features for post-icu mortality prediction. In: AMIA Annual Symposium Proceedings. vol. 2016. American Medical Informatics Association; 2016. p. 827.

25. Kansagara D, Englander H, Salanitro A, Kagen D, Theobald C, Freeman M, et al. Risk prediction models for hospital readmission: a systematic review. Jama. 2011;306(15):1688–1698.

26. Campbell AJ, Cook JA, Adey G, Cuthbertson BH. Predicting death and readmission after intensive care discharge. British journal of anaesthesia. 2008;100(5):656–662.

27. Futoma J, Morris J, Lucas J. A comparison of models for predicting early hospital readmissions. Journal of biomedical informatics. 2015;56:229–238.

28. Liu V, Kipnis P, Gould MK, Escobar GJ. Length of stay predictions: improvements through the use of automated laboratory and comorbidity variables. Medical care. 2010;p. 739–744.

29. Hackbarth G, Reischauer R, Miller M. Report to the Congress: promoting greater efficiency in Medicare. Washington, DC: MedPAC. 2007;.

30. Le Q, Mikolov T. Distributed representations of sentences and documents. In: International conference on machine learning; 2014. p. 1188–1196.

31. Rehurek R, Sojka P. Software framework for topic modelling with large corpora. In: In Proceedings of the LREC 2010 Workshop on New Challenges for NLP Frameworks. Citeseer; 2010.

32. Pedregosa F, Varoquaux G, Gramfort A, Michel V, Thirion B, Grisel O, et al. Scikit-learn: Machine learning in Python. Journal of machine learning research. 2011;12(Oct):2825–2830.

33. Paszke A, Gross S, Massa F, Lerer A, Bradbury J, Chanan G, et al. PyTorch: An imperative style, high-performance deep learning library. In: Advances in Neural Information Processing Systems; 2019. p. 8024–8035.

